# Young people with all forms of shoulder instability demonstrate differences in their movement and muscle activity patterns when compared to age- and sex-matched controls

**DOI:** 10.1101/2023.07.15.23292602

**Authors:** Martin Seyres, Neil Postans, Robert Freeman, Anand Pandyan, Ed Chadwick, Fraser Philp

## Abstract

**Background:** Shoulder-instability is a complex impairment and identifying biomarkers which differentiate subgroups is challenging. Robust methods of measurement and movement protocols for improving our current understanding of muscle activity mechanisms, which may inform subgrouping and treatment allocation are needed.

**Hypothesis:** Null hypothesis: there are no differences between the movement and muscle activity of young-people with shoulder instability (irrespective of aetiology) and age- and sex-matched controls (two-tailed).

**Methods:** Young-people between eight to 18 years were recruited into two groups of shoulder-instability (SI) or and age- and sex-matched controls (CG). All forms of SI were included and young-people with co-existing neurological pathologies or deficits were excluded. Participants attended a single session and carried out four unweighted and three weighted tasks in which their movements and muscle activity was measured using 3D-movement analysis and surface electromyography. Statistical parametric mapping was used to identify between group differences.

**Results:** Data was collected for 30 young-people (15 SI (6M:9F) and 15 CG (8M:7F)). The SI cohort had mean (SD) age, height and weight values of 13.9 years (2.9), 163.0 cm (15.7) and 56.6 kg (17.5) respectively. The CG had age, height and weight values of 13.3 years (3.1), 160.6 cm (16.8) and 52.4 kg (15.1) respectively. The SI group demonstrated consistently more protracted and elevated sternoclavicular joint positions during all movements. Normalised muscle activity in Latissimus dorsi had the most statistically significant between group differences across all movements. The SI group also had increased normalised activity of their middle trapezius, posterior deltoid and biceps muscles whilst activity of their latissimus dorsi, triceps and anterior deltoid were comparatively decreased.

**Discussion:** Young-people with SI may constrain their movements to minimise glenohumeral joint instability. This was demonstrated by reduced variability in joint angles, adoption of different movement strategies across the sternoclavicular and acromioclavicular joints and increased activity of the scapular stabilising muscles, despite achieving similar arm positions to the CG.

**Conclusion:** Young-people with shoulder instability have consistent differences in their muscle activity and movement patterns. Consistently observed differences at the shoulder girdle included increased sternoclavicular protraction and elevation accompanied by increased normalised activity of the posterior scapula stabilising muscles and decreased activity of the posterior humeral mobilising muscles. Existing methods of measurement may be used to inform clinical decision making, however, further work is needed evaluate the prognostic and clinical utility of derived 3D and sEMG data for informing decision making within shoulder instability and associated subgroups.

## Introduction

Shoulder instability is a complex impairment which manifests as excessive translation between the humerus and glenoid resulting in partial subluxation or complete dislocation of the glenohumeral joint. A plethora of classification systems exist which seek to identify pathophysiological mechanisms that are causal or contributory to the presentation of shoulder instability. Broadly these include injury mechanism (traumatic or atraumatic), instability direction, frequency and severity (subluxation/dislocation), and role of body structures and functions (bony morphology, supporting capsular and ligamentous structures and “muscle patterning”) ^17; 22; 23;28^. Socio-economic and personal/social factors may be associated with the impairment but are not explicitly identified or explained in existing models ^23; 28^. However, identification of the most significant factors is important for improving patient outcomes through timely assessment, referral and appropriate treatment allocation.

Considerable emphasis has been placed on the role of the shoulder muscles and their activity profiles, often referred to as “muscle patterning”, in both the diagnosis and rehabilitation of shoulder instability ^28^. Existing phenotypes and classifications, which inform treatment decisions, are based on assumptions which have not been fully validated. From a diagnostic perspective it is hypothesised that different activation profiles or inappropriate muscle co-ordination may cause or contribute to atraumatic instability. Existing studies make assumptions and possibly conflate kinematics, measured surface electromyography (sEMG) and the magnitudes of associated muscle forces, despite not measuring actual muscle force ^30; 33; 35; 44^. Whilst it has been suggested that these differences are causative of shoulder instability, it is also possible and likely that these differences are reflective of the neuromusculoskeletal system adapting and optimising for the constraint of stability in light of underlying congenital, developmental or acquired bony morphology and soft tissue ligamentous changes which may be static or dynamic. It is worth noting that studies linking muscle activity patterns to instability have no normative data prior to the development of instability, have not measured these physiological changes longitudinally or evaluated a comprehensive set of the upper limb muscles and movements ^18; 34^. Any inferences regarding causal mechanisms from these studies should therefore be undertaken with this understanding.

Treatment and rehabilitation of shoulder instability are also influenced by current but incomplete understandings of motor control and muscle structure and function ^40; 47^. Rehabilitation may be aimed at either restoring assumed “normal” or retraining alternative or “optimal” muscle patterning to ensure stability of the joint. In the absence of high-quality or good first principle measurement studies, several treatment philosophies have developed which seek to make use of the “kinetic chain”, “activating the cuff”, co-contraction or redundancy principles ^30; 40^. These assume that the movement strategy being developed to maintain joint stability under one set of task and environmental constraints is retained and transferrable to different task and environmental constraints outside of the rehabilitation context. This is yet to be demonstrated and although it may seem intuitive, they remain conceptual, lacking specificity and are often complicated by imprecise terms and biomechanical principles ^39; 40^.

A biopsychosocial approach, founded on a physiologically accurate and scientifically valid understanding of all mechanisms which contribute to shoulder instability in young people is required for improving diagnostic processes and patient outcomes. Children who present with instability and an unclear mechanism are known to be complex and highly variable, possibly as the developing adolescent system is in an ongoing process of learning and adaptation to evolving maturation related changes ^29; 31; 34^. Whilst imaging modalities such as MR arthrogram and x-ray have given more definitive answers regarding structural features that may contribute to instability, there is no equivalent method for measuring or referencing muscle activity profiles in young people with shoulder instability ^34; 38^. Three-dimensional movement analysis of the lower limb, which includes surface electromyography (sEMG), is used routinely in clinical practice to inform decision making in complex patient groups with disordered control, but has not been widely integrated into clinical upper-limb services due to limited reference protocols and tasks ^41^. There is therefore a need to develop robust methods of measurement and associated movement protocols to improve our current understanding of muscle activity mechanisms in shoulder instability which can be translated into clinical practice. The aim of this study was to identify if there are any movement and muscle activity differences between young-people with shoulder instability and age- and sex-matched controls and quantify these differences where they exist. Our hypothesis was non-directional, with the null hypothesis being that there are no differences between the movement and muscle activity of young people with shoulder instability, irrespective of aetiology (SI), and age- and sex-matched controls (CG).

## Materials and Methods

Ethical approval for this study was gained from West Midlands - South Birmingham Research Ethics Committee REF:20/WM/0021. This trial is registered on ClinicalTrials.gov Identifier: NCT04267354 available at: https://clinicaltrials.gov/ct2/show/NCT04311216.

### Study design

This study recruited participants from two different sampling frames. These were a group of young people with shoulder instability (SI) and an age- and sex-matched control group (CG). Participants were recruited from a single tertiary centre and the study was advertised across regional clinical centres and social media. A total of five additional centres signposted participants to the study. Recruitment was over a 24-month period. The overall recruitment rate was 81% with seven out of 37 participants approached declining or unable to take part in the study.

Following informed consent to participate in the study, all participants attended a single measurement session for demographic, clinical and 3D-movement assessment of their upper-limb. Participants were provided with paper diaries to record their instability episodes and followed up on monthly basis for one year using phone calls and electronic communications to record any episodes of instability. This paper describes the baseline biomechanical measurements and identified movement and muscle activity differences between groups.

### Inclusion criteria

For both groups, young people aged between eight and 18 were included unless there were any co-existing neurological pathologies or deficits. For the SI group they were included if they had symptomatic instability with at least one sign of instability on clinical examination during the sulcus, apprehension or anterior and posterior shift load tests. This included patients with all forms of instability i.e. recurrent, first-time, multidirectional, atraumatic and traumatic instability and those who had instability following previous surgery.

### Exclusion criteria

For the SI group they were excluded if they were previously surgically managed and did not have any further episodes of instability following the intervention. For the CG they were excluded if they had any previous presentation to a health care professional with a diagnosis of shoulder instability, a shoulder injury within the last three months on the arm being assessed that had not resolved, previous surgical intervention on the arm being assessed or ongoing or pending medical management, diagnostic investigations or rehabilitation on the arm being assessed.

### Demographic and clinical assessments

Clinical assessments included recording of the following instability features: type, (single episode or recurrent), apprehension, guarding or laxity in the sulcus, anterior and posterior shift load, and apprehension relocation test, as well as Beighton score of hypermobility and grip strength testing. Additional questions included relevant past medical history, time since last instability episode, side(s) of instability, self-reported dislocation or subluxation, direction and number of subluxation or dislocation episodes.

### 3D movement analysis measurement protocol

An overview of the marker cluster and sEMG placement for data collection is shown in Figure 1. Retroreflective marker clusters were placed on the thorax, acromion, humerus, forearm and hand segments adapted from Jaspers et al ^19; 20^ and available at available at https://datacat.liverpool.ac.uk/. sEMG electrodes were placed on the middle trapezius, infraspinatus, triceps, latissimus-dorsi, deltoid (posterior and anterior), pectoralis-major, biceps, wrist-flexor and extensor muscles according to SENIAM guidelines ^12^ and Criswell et al ^8^. For subject calibration, the Pellenburg wand was used for virtual marker identification of the following bony landmarks: C7 spinous process (C7), T8 spinous process (T8), Insicura Jungularis (IJ), Processus Xiphoideus (PX), Articulation Sternoclavicularis (SC), Articulation Acromioclavicularis (AC), Processus Coracoideus (PC), Trigonum Scapulae (TS), Angulus Inferior (AI), Angulus Acromialis (AA), Lateral Epicondyle (LE), Medial Epicondyle (ME), Radial Styloid (RS), Ulnar Styloid (US), Styloid process of 3rd Metacarpal (MC3) and distal heads of the 2^nd^, 3^rd^ and 5^th^ metacarpophalangeal joints (MCP2, MCP3 and MCP5) ^19–21^.

**Figure 1.** Overview of marker clusters and EMG placement in study.

Participants’ movements were assessed in four unweighted movements (flexion, abduction, abduction to 45° with axial rotation, and hand to back of head) and three self-selected weighted tasks of 0.5kg, 1.0kg or 1.5kg (flexion, abduction, abduction to 45° with axial rotation) in that order. Participants were initially shown the movements by the assessor and then asked to carry them out to a count of 4 seconds up, 4 seconds down, mirroring the assessor who was positioned in front of them. Data were collected at 100Hz using a Vicon motion capture system (12 V5-Vantage motion analysis cameras, two synchronous coronal and sagittal video recordings and Delsys Trigno electromyography system sampling at 2000Hz). Interpolation for any missing marker data was performed as appropriate using rigid body, pattern and spline filling pipelines available within Vicon Nexus 2.12.1 ^1^.

### Data processing and analysis

Joint angles were calculated using inverse kinematics and the Wu shoulder model ^54^ in Opensim 4.4 ^9;45^. Definitions of joint co-ordinate systems were consistent with International Society of Biomechanics (ISB) recommendations ^53^. Model scaling and evaluation were consistent with best practice frameworks in which a scaling ratio for each bone was estimated from selected marker pairs for each segment, obtained during the anatomical marker identification for static calibration (Appendix 1). ^13;46^. Kinematics were smoothed using a Savitzky-Golay filter, with a window size of 99 and a polynomial order of two ^54^. The filter and parameters were selected as they perform well when during high-frequency acceleration-time signals when compared to alternative methods, and based on our data set, performed the best for removal of noise whilst preserving the underlying signal ^43^.

The glenohumeral joint origin was determined through geometrical scaling. This method was selected over regression, functional or offset methods as the presence of excessive translation (instability) in this cohort would likely violate the assumptions required for implementation of the aforementioned methods. To reflect the angles observed by clinicians in practice, thoracohumeral and scapulothoracic angles were calculated for positions of the arm and scapula with respect to the thorax. Additionally, joint-specific angles for the glenohumeral, sternoclavicular and acromioclavicular joints were also calculated.

sEMG signals were band-pass filtered between 10-400 Hz using a second order Butterworth filter, and zero lag correction offset was then applied ^52^. sEMG was normalised to the maximum encountered activation across any of the movement activities, including isolated movements against resistance for quality control, grip, weighted and unweighted tasks. No maximum voluntary contraction (MVC) testing was carried out to minimise risk of further instability during data collection and as this is known to be highly variable, particularly in pathological populations ^48^.

Group demographics are presented as frequencies. Statistical parametric mapping (SPM) with a Student’s t-test was used to identify between group differences for joint kinematics and normalised sEMG signals ^37^. SPM with Student t-test was used as the aim of our study was to evaluate if there are differences between two groups at the level of the impairment rather than on the basis of a theoretical classification system or aetiological subgroups.

For between group comparisons, thoracohumeral and thoracoscapular angles were reported, to reflect clinician’s observation in practice, but were not included in the statistical analysis given that they are not physiologically representative and compliant with ISB recommendations or generated in the selected model ^53; 54^. In order to identify kinematic differences that could be considered clinically meaningful a threshold of ≥ 10° was identified. To identify the variability associated with our protocol, mean and 95% CI were reported for the CG kinematics and the threshold of ≥ 10° for 95% CI ranges was selected to identify joint planes of movement considered has having considerable movement variability. A threshold of ≥ 10° was also selected for between SI and CG group differences, as differences of this magnitude are likely apparent with clinical observation and larger than the error of measurement and variability associated with our methodologies ^42^. C3D files used for 3D movement and sEMG analysis are available at https://datacat.liverpool.ac.uk/.

## Results

### Group demographics

Data were collected for 30 young people, 15 with shoulder instability (SI) and 15 sex- and age-matched controls (CG) with demographic data presented in Table 1.

**Table 1.** Participant demographics for all study participants.

|  | **CG** | **SI** |
| --- | --- | --- |
| Age (years) | 13.3 (3.1) | 13.9 (2.9) |
| Height (cm) | 160.6 (16.8) | 163.0 (15.7) |
| Weight (kg) | 52.4 (15.1) | 56.6 (17.5) |
| Male to Female (M:F) | 8:7 | 6:9 |
| Beighton score (median (IQR)) | 2 (0.5 to 2.5) | 6 (2 to 6.5)** |
| Grip strength Mean max value left (kgf) | 28 (12.5) | 26.7 (10.5) |
| Grip strength Mean max value right (kgf) | 31.2 (13.6) | 28.9 (10.4) |
| Dominant hand (L:R) | (0:15) | (1:14) |
| Number of participants whose non-dominant hand was assessed for 3D | 3 (L) | 5(L)* |
| Instability side (bilateral:left:right) | N/A | (10:1:4) |
| Side assessed in 3D movement (L:R)* | (3:12) | (6:9) |
| Weight selection for loaded tasks (0.5kg:1.0kg:1.5kg) | (1:3:11) | (1:5:9) |

\* discrepancy due to drop outs for the side
\*\* one participant unable to do 5<sup>th</sup> digit (little) fingers due to previous injuries

### Shoulder instability group

For the SI group, three participants presented for data collection having sustained a first-time episode of shoulder instability and 12 after recurrent episodes of instability. The most common form of instability experienced prior to attendance was subluxation, reported by 13 participants. Only one participant reported having experienced a definite dislocation and one participant was unsure if the most recent episode was a subluxation or dislocation. Ten participants had an atraumatic aetiology, four reported a traumatic aetiology, and one reported an ambiguous overlapping atraumatic/ traumatic aetiology. Two participants were unable to identify the direction of their instability. Subjective reports of anterior instability were reported by seven participants, two reported posterior instability, two reported inferior instability and two reported multidirectional instability in the posterior/inferior and anterior/inferior directions.

Length of time since last instability episode ranged from 4 hours to 32 weeks with a mean time of 7 weeks (SD 9 weeks). Two participants were unable to recall the length of time since their last episode. The number of self-reported subluxations ranged from one to more than 180 and the number of self-reported dislocations ranged from one to more than 90, with some participants and parents estimating the total number (subluxations and dislocations) by the product of the length of time since the onset of instability and a conservative daily frequency for instability episodes in cases of difficulties in recalling exact numbers.

### Relevant past medical history

Two participants had formal diagnosis of connective tissue or hypermobility disorders. Of these one had an atraumatic aetiology and one had an ambiguous overlapping atraumatic/ traumatic mechanism.

## Joint kinematics

### Age and sex-matched controls (CG)

To provide normative reference values and identify the variability associated with our protocol, mean and 95% CI are presented for the CG only in Table 2.

**Table 2.**
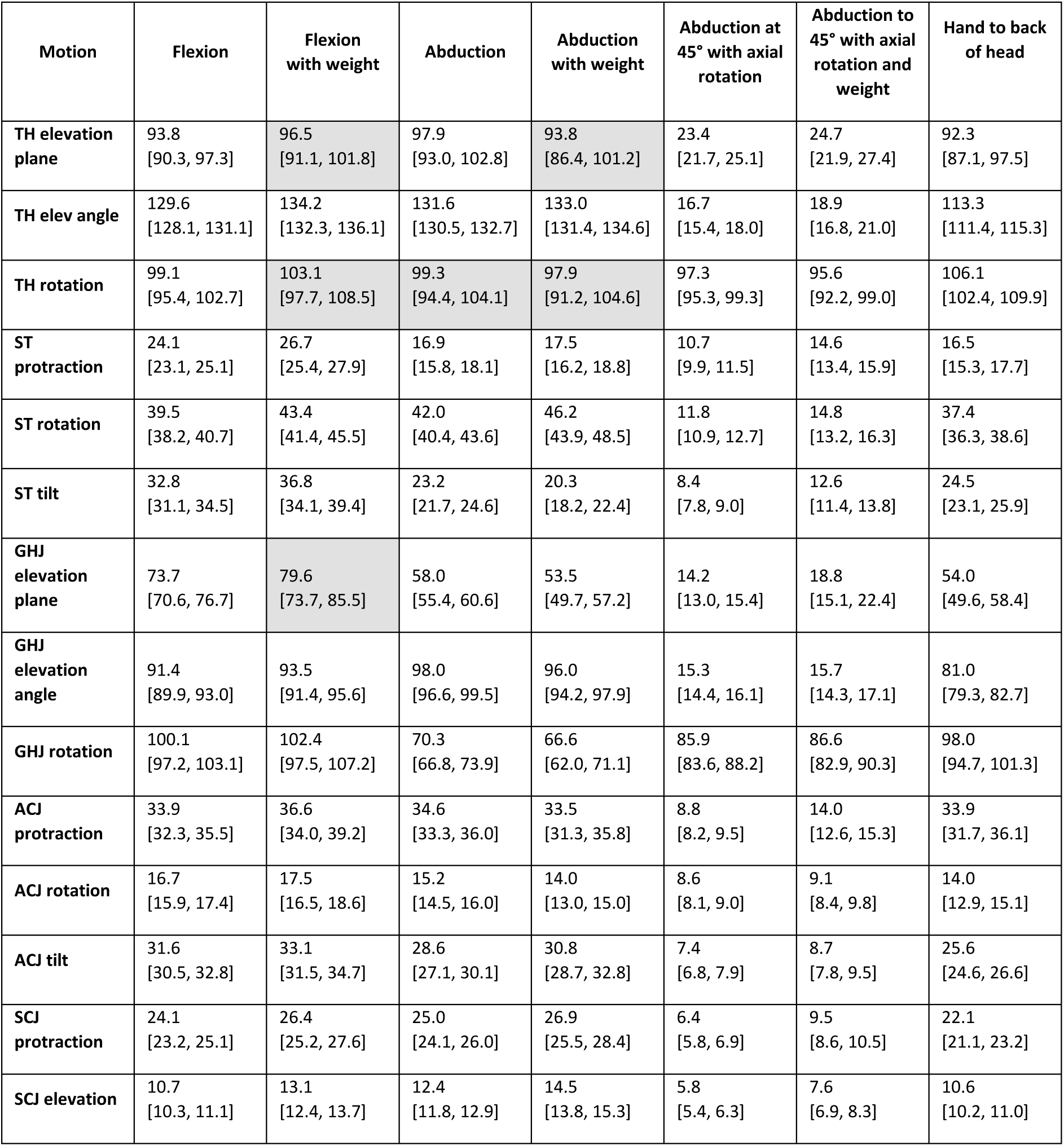
Mean ROM [95% CI] values for all joints and movements in the CG (degrees) *Shaded boxes indicate 95% CI ranges ≥ 10 degrees ; TH = thoracohumeral, ST = scapulothoracic, GHJ = glenohumeral joint, ACJ = acromioclavicular joint, SCJ = sternoclavicular joint*

Overall the majority of movements had 95% CI ranges that did not exceed more than 10°. The widest 95% CI were seen in the weighted abduction movement for the planes of thoracohumeral elevation and rotation. Three participants experienced episodes of shoulder instability (subluxations) during the abduction at 45° with axial rotation, weighted and unweighted tasks.

Mean Range of Motion values and standard deviations (SD) for all joint planes of movement and associated tasks are presented in Table 3 for both groups.

**Table 3.**
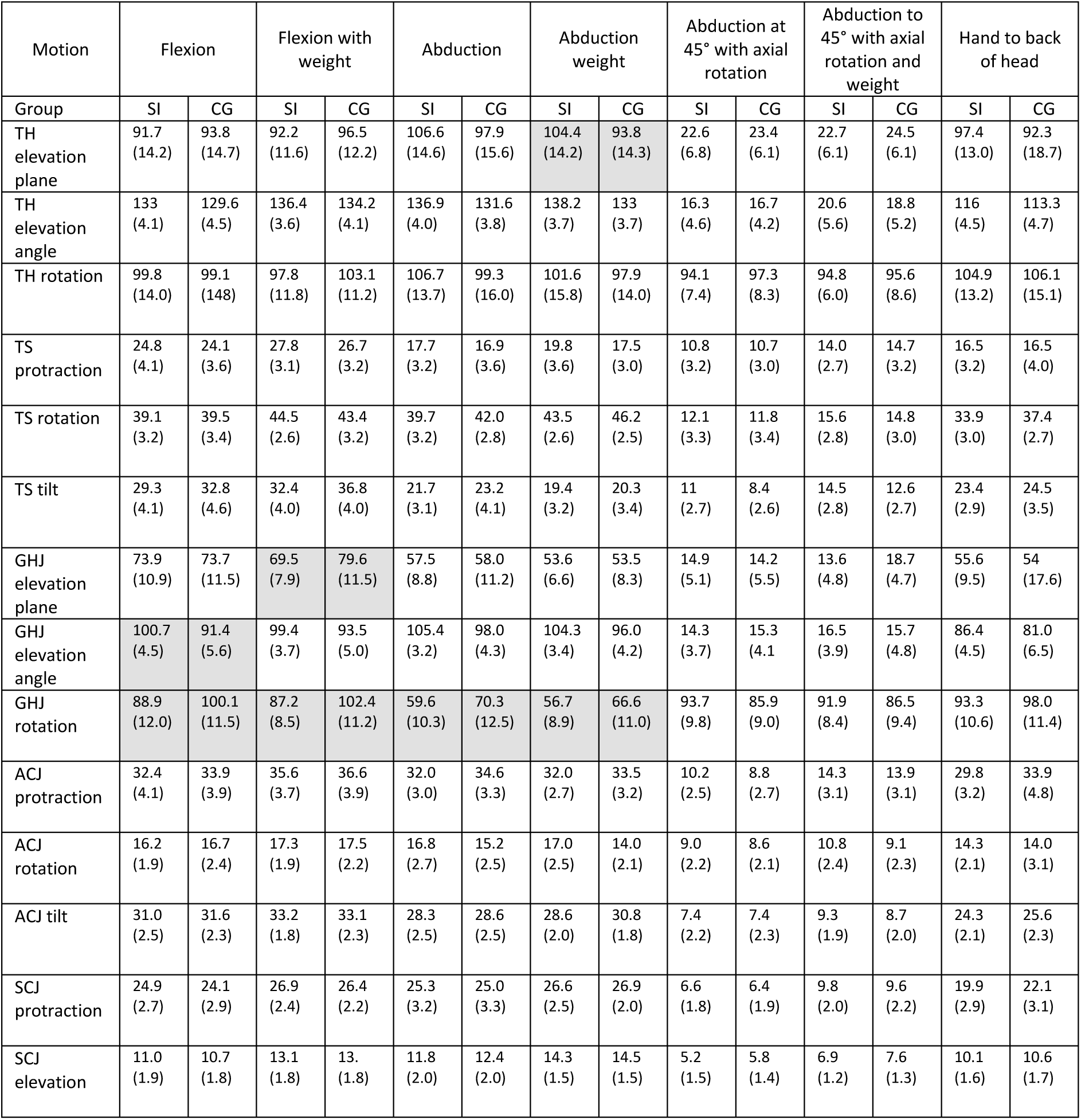
ROM values for planes of movement across all joints and movement tasks for the SI and CG (degrees) *Shaded boxes indicate between group differences ≥ 10 degrees; TH = thoracohumeral, ST = scapulothoracic, GHJ = glenohumeral joint, ACJ = acromioclavicular joint, SCJ = sternoclavicular joint*

Mean between group differences of 10° or more were observed most frequently in the Glenohumeral rotation plane for the movements of both weighted and unweighted flexion and abduction. Statistically significant between group differences for kinematics in SI and CG are reported in Figure 2.

**Figure 2.**
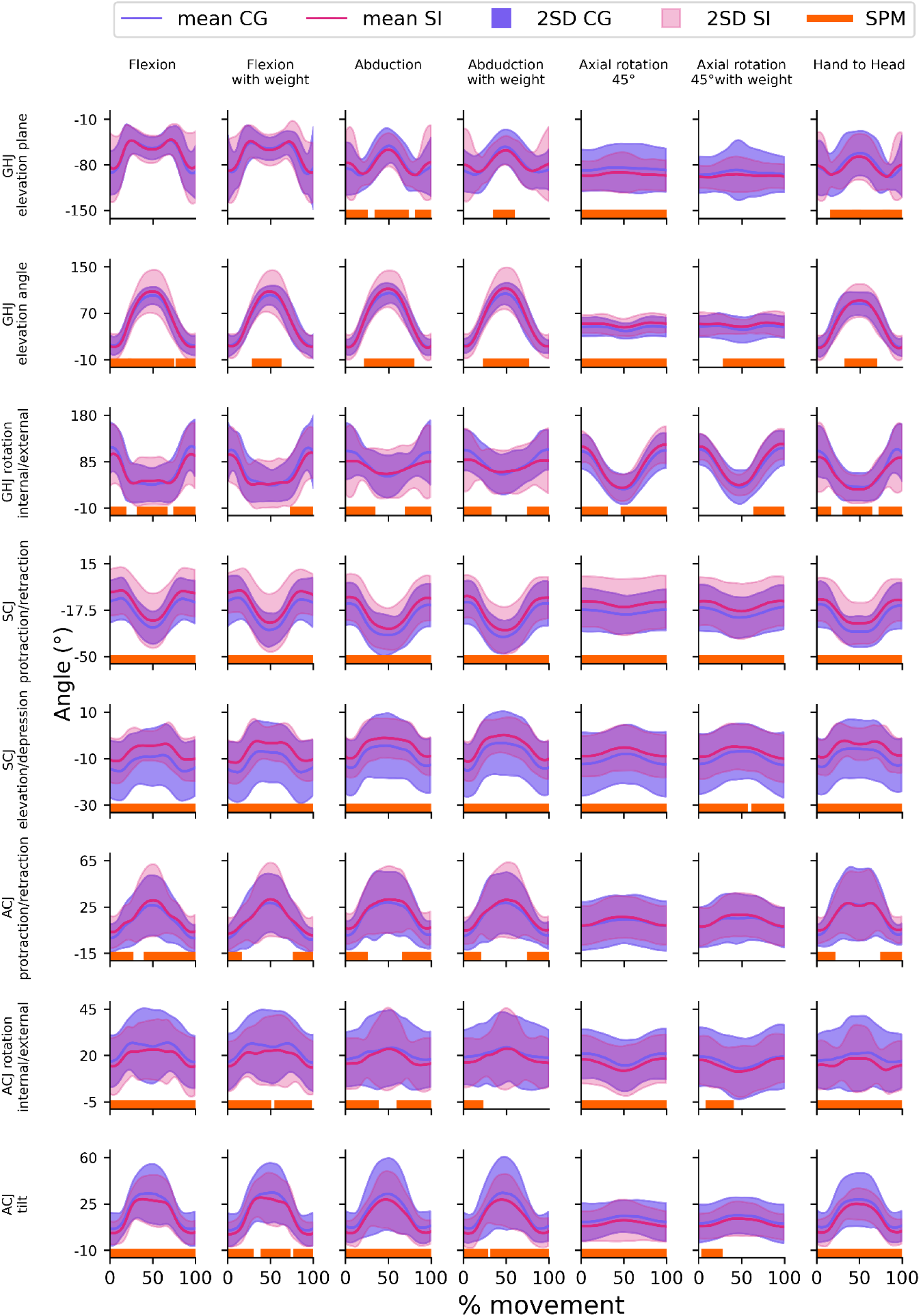
SI and CG kinematics for all movements and SPM. Joint angles for all joints and all movements. Lines show mean group angles, shaded areas indicate the 2SD, and the orange bars on the horizontal axis highlight regions of statistically significant difference between group using statistical parametric mapping (SPM). Column headings Flexion, Flexion with weight, Abduction, Abduction with weight, Axial rotation 45° = Abduction at 45° with axial rotation, Axial rotation 45° = Abduction to 45° with axial rotation and weight, Hand to head = Hand to back of head. GHJ = glenohumeral joint, ACJ = acromioclavicular joint, SCJ = sternoclavicular joint.

Statistically significant between group differences were observed across almost all movement tasks and for all joint planes of movement. Consistent differences across the entire movement cycle and for all movement tasks were observed in the sternoclavicular protraction/retraction and elevation/ depression planes. The SI group adopted a more protracted and elevated sternoclavicular joint during all movements. In most movements this was accompanied by less internal rotation and upwards tilt at the acromioclavicular joint. No differences were observed in the unweighted and weighted flexion tasks for the glenohumeral joint plane of elevation, and unweighted and weighted abduction to 45° with axial rotation acromioclavicular joint protraction/retraction plane. Statistically significant between group differences for measured sEMG in SI and CG are reported in Figure 3.

**Figure 3.**
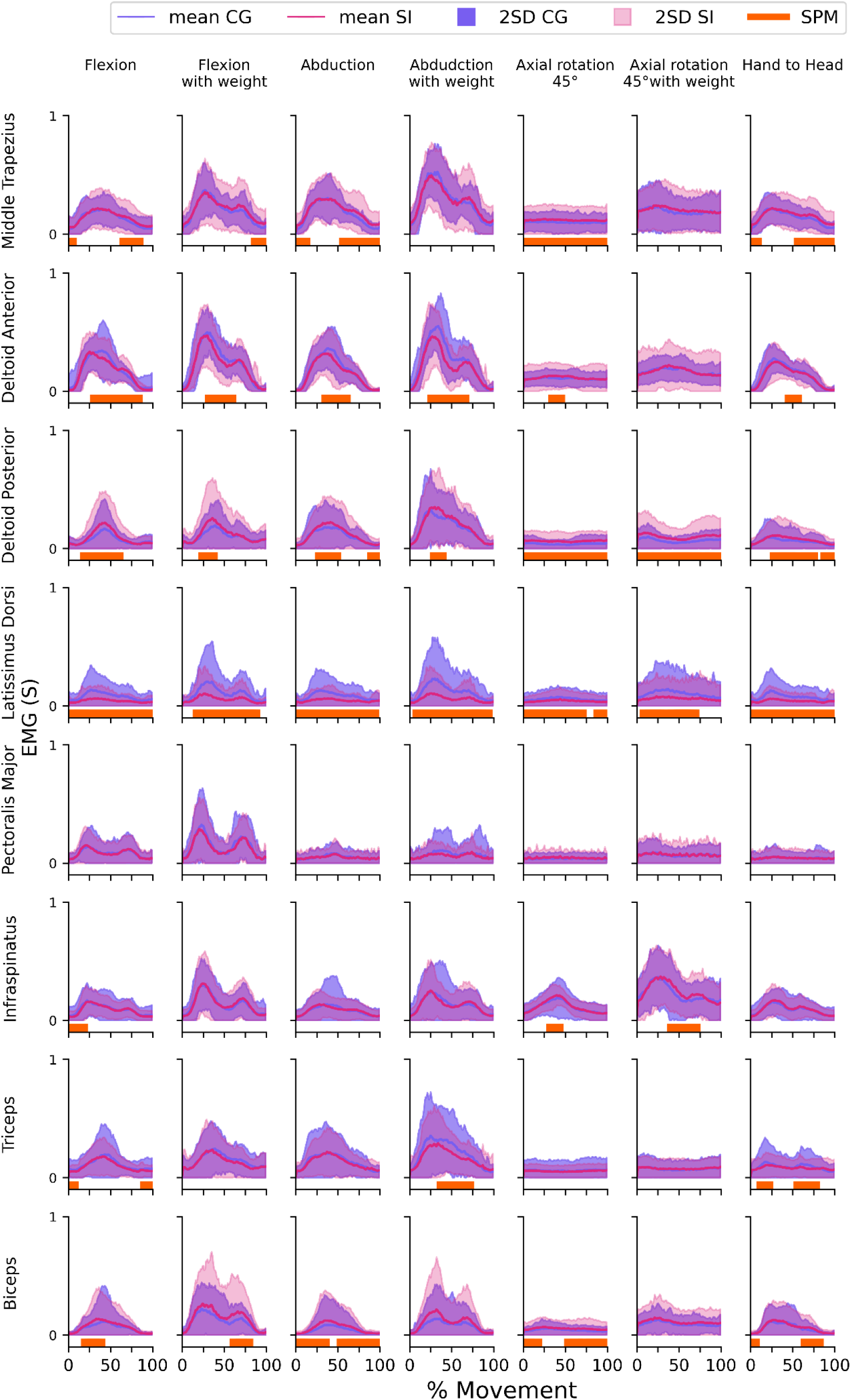
SI and CG sEMG for all movements and SPM. Muscle activity profiles for all muscles and all movements. Lines show mean group angles, shaded areas indicate the 2SD, and the orange bars on the horizontal axis highlight regions of statistically significant difference between group using statistical parametric mapping (SPM). Column headings Flexion, Flexion with weight, Abduction, Abduction with weight, Axial rotation 45° = Abduction at 45° with axial rotation, Axial rotation 45° = Abduction to 45° with axial rotation and weight, Hand to head = Hand to back of head. GHJ = glenohumeral joint, ACJ = acromioclavicular joint, SCJ = sternoclavicular joint.

No statistically significant between group differences were observed in the Pectoralis major muscle in any of the movement tasks. Latissimus dorsi showed significant differences across a greater proportion of tasks. Weighted tasks had fewer muscle activity differences identified as being statistically significant between groups than unweighted tasks. When compared to the CG, the SI group had increased normalised activity of their middle trapezius, posterior deltoid and biceps muscles whilst activity of their latissimus dorsi, triceps and anterior deltoid were comparatively decreased. It appears that muscles which control scapular movement on the posterior compartment of the body (middle trapezius, posterior deltoid and biceps) have higher normalised activity whilst muscles that primarily control humeral movement have lower normalised activity (latissimus dorsi and triceps). However, the inverse is true for muscles on the anterior portion of the body with increased biceps and decreased anterior deltoid activity.

## Discussion

The aim of this study was to identify if there are any movement and muscle activity differences between young-people with shoulder instability and age- and sex-matched controls and quantify these differences where they exist. Fundamental research evaluating mechanisms for shoulder instability in young people is very limited and our cohort is one of the youngest evaluated ^4; 18; 36; 50^. Our study provides evidence that following an episode of instability, there are muscle activity and movement pattern differences between those with shoulder instability and age- and sex-matched controls. Using the protocol developed it been possible to quantify the variability in upper-limb movements and this can be used to identify meaningful differences or changes in an age- and sex-matched sample with shoulder instability. It has also been possible to identify between group differences, considered statistically significant, across several muscles, joint planes of movement and phases in the movement cycle.

Overall thoracohumeral angles and by proxy, arm positions, during the movements were similar between groups, however, the SI group adopted different movement strategies across the shoulder girdle joints to achieve this, mainly at the sternoclavicular and acromioclavicular joints. The joint planes of movement and periods of the movement cycle identified as having statistically significant differences varied according to the movement being carried out, although some behaviours were common to the SI group. Consistent differences were seen for all movements and across the entire movement cycle at the sternoclavicular joint, with the SI group adopting a more protracted and elevated sternoclavicular joint during the movements. In most movements this was accompanied by less internal rotation and upwards tilt at the acromioclavicular joint. Common differences in behaviour were also seen in the sEMG measurements of the SI group. Direct comparison of our findings to other studies is challenging due to variations in the movements conducted, number and selection of segments and muscles measured, and methods of analysis used for joint kinematics and sEMG. However, where methods are generally comparable, our results are similar for the anterior deltoid, infraspinatus and triceps muscles which had higher normalised activity profiles in the SI group ^15; 16; 49;50^. Biceps was identified as having higher levels of normalised activity in the SI group which is different to other published studies which reported lower levels of normalised activity ^15; 16; 50^. Differences in our results may reflect the fact that whilst movements between studies were broadly similar, they were not identical. As a result of the impairment, the observed movement and muscle activity patterns of those with shoulder instability may be constraining movements around the shoulder girdle to maximise stability of the glenohumeral joint.

Pectoralis major activity was not identified as being statistically significantly different between groups for any of the movements assessed. This may be unexpected as it is often assumed to be a driver for anterior and possibly multidirectional instability given its action on the humerus. Whilst pectoralis major was not identified as being different in the combined group data, there was an individual with instability who did present with a different profile of pectoralis activation during some of the measurements. Instability may occur under different task or environmental constraints not evaluated in our study and highlights the challenges of developing a universal protocol for assessing the upper-limb. Further work will be needed to explore how the testing protocol can be extended to match with different subgroups or pathophysiological presentations. Future research may customise protocols on the basis of clinical signs or a form of baseline screening. Furthermore, our study aimed to identify differences related to instability at a group rather than an individual level and the overall number of instability episodes within a movement task were also relatively low compared to the overall number of repetitions. Whilst these methods of measurement can be used to inform clinical decision making on an individual basis, further work is needed evaluate the prognostic and clinical utility of derived 3D and sEMG data for informing decision making within shoulder instability ^3; 25; 39; 41; 46^.

Within existing instability classification systems and accompanying treatment philosophies, it is not clear if the observed movements and muscle activity develop in response to the impairment or are a significant contributing factor to its development ^11; 28; 40^. Unpicking the relevance of the identified muscle activity profiles and movement patterns is challenging in young-people given that their neuromusculoskeletal system is continually developing alongside possible changes to their environment (home and school life) and personal factors ^51^. These developments are often overlaid with changes to body structure, body functions and personal factors that contribute to the impairment of instability ^51^. Furthermore, there is limited longitudinal natural history 3D kinematic and muscle activity data and an absence of comparative data pre and post the occurrence of an initial instability episode. We propose that the observed differences are resultant from the SI group optimising their muscle activity and movement patterns for stability in response to any underlying changes in their perception, bony or soft tissue structures in their shoulder ^47^. When comparing the weighted and unweighted tasks there were fewer differences between the SI and CG groups for both kinematics and sEMG measures during the weighted tasks. Under loaded or novel conditions, the CG may also have constrained their movements for stability. Therefore, it appears than in young-people whose joint stability is challenged they will constrain movements around the shoulder girdle but may transition to more variable movement patterns as their ability to maintain stability is improved, subsequently increasing the degrees of freedom available and utilising the passive forces of the soft tissues ^47^. This is consistent with existing motor control paradigms and experimental studies but further work investigating indices of stability are needed to evaluate this ^2; 32; 47^. This has implications for rehabilitation as it demonstrates there is no universally ideal or normative movement pattern and clinicians should avoid trying to impose assumed ‘best-movement patterns’ on those undergoing rehabilitation. It may also explain why existing treatment approaches which integrate early weightbearing or loading have positive results. The applied load may constrain the task, effectively reducing the degrees of freedom and requiring increased muscle co-contraction, naturally leading to increased glenohumeral joint stability ^5; 29^.

Our results demonstrate that assessment of upper-limb movements is complex and that existing methods of clinical assessment may not capture this complexity. Being able to accurately measure the differences and changes observed in a reliable way without technology is unlikely given the large number and magnitude of differences seen within and between movements. Differences in joint planes of movement and muscle activity was dependant on the movement task being carried out. The assessment of a single movement may therefore not be sufficient for identifying links between the impairment of interest and associated biomechanical data needed to inform clinical decision making. This is consistent with studies investigating 3D upper limb function in other populations ^6; 41^. However, within shoulder instability, it is not clear which movement tasks and generated biomechanical data are the most important for informing decision making. Selection of tasks for evaluation with 3D movement analysis and sEMG needs to be considered alongside the large volume of data that is generated using these methods which can limit interpretability and translation into clinical practice.

## Limitations

Whilst our protocol was able to identify differences in the joint movements and muscle activity patterns of those with shoulder instability, this was done in a limited number of movements and superficial muscles. Whilst sEMG does not allow for direct measurement of deeper glenohumeral or scapular stabilising muscles e.g. the rotator cuff group or serratus anterior, sEMG is preferable for use in young-children for ethical and pragmatic reasons given that it is non-invasive with fewer risks. Musculoskeletal modelling tools may be used to approximate information about muscles that are challenging to measure and provide some further understanding of their role ^9; 46^. sEMG can be affected by cross-talk, although appropriate placement according to established guidelines, trained experts and quality control checks as carried out in our study can mitigate against this.

During the protocol participants carried out movements over a large range of motion. Calculation of joint angles, mainly at the glenohumeral joint, at the extremes of motion can result in a number of mathematically correct but clinically counterintuitive solutions given that differentiation of the planes of movement can be challenging. Calculation of joint kinematics was performed using established modelling conventions but interpretation of kinematic results should be carried out with this understanding. In our study we chose to group participants at the level of the impairment as there is limited fundamental science demonstrating proposed mechanisms and existing classification systems are conceptual and can be non-discriminatory or prone to misclassification ^23; 34; 40^. Further subgroup analysis informed by existing classification frameworks and traumatic or atraumatic aetiological causes may be carried out in future work. However, a fundamental step is to ensure that categories are developed on the basis of appropriate measures or first principles and that the underlying pathology is not confused with the impairment ^10^.

Our study only conducted measurements at a single time-point in young-people aged between eight and 18. Further longitudinal measures in a larger sample with a wider range of ages (young people and adults) and aetiological subgroups is required for a robust understanding of factors that contribute to shoulder instability. Future research may also include other biopsychosocial factors relevant to shoulder instability. It is possible that the differences observed between groups may be influenced by the order, number of repetitions and speeds of movement in our protocol i.e. several unweighted movement repetitions progressing to weighted repetitions. Variation in any components of the protocol may potentially result in different outcomes. It is recognised that the weights used in our study were relatively low and individuals may have been working at different levels of their maximum capacity. Given the exploratory nature of the study and ethical considerations to minimise risk of harm, weight selection was a pragmatic choice. Additionally, only participants who were able to engage with the entire measurement protocol were included. Our selected protocol may not be feasible in patients with more severe forms of instability. Despite this our protocol was able to measure the impairment of interest, which usually occurred in the abduction at 45° and axial rotation tasks (weighted and unweighted), a position known to challenge the stability of the glenohumeral joint and occurred towards the end of the movements being assessed.

During the clinical assessment the number of self-reported instability episodes was high when compared to other studies and likely subject to recall bias, evidenced by some participants and their parents being unable to recall a definitive number or features and timelines related to the instability ^14; 26; 27; 31^. Existing studies recognise that the true incidence and prevalence of shoulder instability is likely underreported and the true long-term health and economic impact of recurrent instability, particularly subluxations, is unknown ^7; 24; 27^. Young people classified as having atraumatic instability can experience multiple episodes that do not interfere with overall function and sometimes experience a delayed presentation to healthcare professionals owing to a combination of absence of knowledge regarding their condition and dependency on parents for accessing health services ^14; 26; 27;31^. Further research should evaluate the true economic and healthcare costs for recurrent shoulder instability facilitated by improved methods of long-term follow-up, recording of instability episodes and linked to long-term health outcomes.

## Conclusions

Young people with shoulder instability have consistent differences in their muscle activity and movement patterns when compared to age- and sex-matched controls. Consistently observed differences at the shoulder girdle included increased sternoclavicular protraction and elevation accompanied by increased normalised activity of the posterior scapula stabilising muscles and decreased activity of the posterior humeral mobilising muscles. Young people with shoulder instability demonstrated less variability in their overall movements and are likely constraining their movements to minimise glenohumeral instability. Existing methods of measurement may be used to inform clinical decision making, however further work is needed evaluate the prognostic and clinical utility of derived 3D and sEMG data for informing decision making within shoulder instability and associated subgroups.

## Data Availability

All data produced are available online from the Liverpool research data catalogue https://datacat.liverpool.ac.uk/

## References

©Vicon Motion Systems Ltd. Vicon Nexus. In; 2022.

Ameln DJD, Chadwick EK, Blana D, Murgia A. The Stabilizing Function of Superficial Shoulder Muscles Changes Between Single-Plane Elevation and Reaching Tasks. IEEE transactions on bio-medical engineering 2019;66:564–572. 10.1109/tbme.2018.2850522

Arnold A, Liu M, Ounpuu S, Swartz M, Delp S. The role of estimating hamstrings lengths and velocities in planning treatments for crouch gait. Gait and Posture 2006;23:273–281.

Barden JM, Balyk R, Raso VJ, Moreau M, Bagnall K. Atypical shoulder muscle activation in multidirectional instability. Clinical Neurophysiology 2005;116:1846–1857. https://doi.org/10.1016/j.clinph.2005.04.019

Bateman M, Osborne SE, Smith BE. Physiotherapy treatment for atraumatic recurrent shoulder instability: Updated results of the Derby Shoulder Instability Rehabilitation Programme. Journal of Arthroscopy and Joint Surgery 2019;6:35–41. https://doi.org/10.1016/j.jajs.2019.01.002

Cacioppo M, Loos A, Lempereur M, Brochard S. Bimanual movements in children with cerebral palsy: a systematic review of instrumented assessments. Journal of NeuroEngineering and Rehabilitation 2023;20:26. 10.1186/s12984-023-01150-7

Comadoll SM, Landry Jarvis D, Yancey HB, Graves BR. The financial burden associated with multiple shoulder dislocations and the potential cost savings of surgical stabilization. JSES Int 2020;4:584–586. 10.1016/j.jseint.2020.04.023

Criswell E. Cram’s introduction to surface electromyography: Jones & Bartlett Publishers; 2010. (ISBN No. 1449663621)

Delp SL, Anderson FC, Arnold AS, Loan P, Habib A, John CT et al. OpenSim: open-source software to create and analyze dynamic simulations of movement. IEEE transactions on biomedical engineering 2007;54:1940–1950.

Gough MS, Adam. The Musculoskeletal System in Children with Cerebral Palsy. A philosp[hical approach to management: Mac Keith Press; 2023. (ISBN No. 9781911612537)

Griffin J, Jaggi A, Daniell H, Chester R. A systematic review to compare physiotherapy treatment programmes for atraumatic shoulder instability. Shoulder & Elbow 2022:17585732221080730. 10.1177/17585732221080730

Hermens HJ, Freriks B, Merletti R, Stegeman D, Blok J, Rau G et al. European recommendations for surface electromyography. Roessingh research and development 1999;8:13–54.

Hicks JL, Uchida TK, Seth A, Rajagopal A, Delp SL. Is my model good enough? Best practices for verification and validation of musculoskeletal models and simulations of movement. Journal of biomechanical engineering 2015;137.

Hung NJ, Darevsky DM, Pandya NK. Pediatric and Adolescent Shoulder Instability: Does Insurance Status Predict Delays in Care, Outcomes, and Complication Rate? Orthopaedic journal of sports medicine 2020;8:2325967120959330–2325967120959330. 10.1177/2325967120959330

Illyés Á, Kiss J, Kiss RM. Electromyographic analysis during pull, forward punch, elevation and overhead throw after conservative treatment or capsular shift at patient with multidirectional shoulder joint instability. Journal of Electromyography and Kinesiology 2009;19:e438–e447.

Illyés A, Kiss RM. Electromyographic analysis in patients with multidirectional shoulder instability during pull, forward punch, elevation and overhead throw. Knee Surgery, Sports Traumatology, Arthroscopy 2007;15:624–631.

Jaggi A, Herbert RD, Alexander S, Majed A, Butt D, Higgs D et al. Arthroscopic capsular shift surgery in patients with atraumatic shoulder joint instability: a randomised, placebo-controlled trial. British Journal of Sports Medicine 2023:bjsports-2022-106596. 10.1136/bjsports-2022-106596

Jaggi A, Noorani A, Malone A, Cowan J, Lambert S, Bayley I. Muscle activation patterns in patients with recurrent shoulder instability. International journal of shoulder surgery 2012;6:101.

Jaspers E, Feys H, Bruyninckx H, Cutti A, Harlaar J, Molenaers G et al. The reliability of upper limb kinematics in children with hemiplegic cerebral palsy. Gait & Posture 2011;33:568–575. https://doi.org/10.1016/j.gaitpost.2011.01.011

Jaspers E, Feys H, Bruyninckx H, Harlaar J, Molenaers G, Desloovere K. Upper limb kinematics: development and reliability of a clinical protocol for children. Gait Posture 2011;33:279–285. 10.1016/j.gaitpost.2010.11.021

Jaspers E, Monari D, Molenaers G, Feys H, Desloovere K. ULEMA—upper limb evaluation in movement analysis: open source custom made MATLAB based software. Gait & Posture 2014:S76–S77.

Kuhn JE. A new classification system for shoulder instability. British Journal of Sports Medicine 2010;44:341. 10.1136/bjsm.2009.071183

Kuhn JE, Helmer TT, Dunn WR, Throckmorton VT. Development and reliability testing of the frequency, etiology, direction, and severity (FEDS) system for classifying glenohumeral instability. J Shoulder Elbow Surg 2011;20:548–556. 10.1016/j.jse.2010.10.027

Kuroda S, Sumiyoshi T, Moriishi J, Maruta K, Ishige N. The natural course of atraumatic shoulder instability. Journal of Shoulder and Elbow Surgery 2001;10:100–104. https://doi.org/10.1067/mse.2001.111962

Laracca E, Stewart C, Postans N, Roberts A. The effects of surgical lengthening of hamstring muscles in children with cerebral palsy–the consequences of pre-operative muscle length measurement. Gait & posture 2014;39:847–851.

Lawton RL, Choudhury S, Mansat P, Cofield RH, Stans AA. Pediatric shoulder instability: presentation, findings, treatment, and outcomes. Journal of Pediatric Orthopaedics 2002;22:52–61.

Leroux T, Ogilvie-Harris D, Veillette C, Chahal J, Dwyer T, Khoshbin A et al. The epidemiology of primary anterior shoulder dislocations in patients aged 10 to 16 years. Am J Sports Med 2015;43:2111–2117. 10.1177/0363546515591996

Lewis A, Kitamura T, Bayley J. (ii) The classification of shoulder instability: new light through old windows! Current Orthopaedics 2004;18:97–108.

Liaghat B, Skou ST, Søndergaard J, Boyle E, Søgaard K, Juul-Kristensen B. Short-term effectiveness of high-load compared with low-load strengthening exercise on self-reported function in patients with hypermobile shoulders: a randomised controlled trial. Br J Sports Med 2022;56:1269–1276. 10.1136/bjsports-2021-105223

Lluch-Girbés E, Requejo-Salinas N, Fernández-Matías R, Revert E, Vila Mejías M, Rezende Camargo P et al. Kinetic chain revisited: Consensus expert opinion on terminology, clinical reasoning, examination and treatment in people with shoulder pain. J Shoulder Elbow Surg 2023. 10.1016/j.jse.2023.01.018

Longo UG, van der Linde JA, Loppini M, Coco V, Poolman RW, Denaro V. Surgical Versus Nonoperative Treatment in Patients Up to 18 Years Old With Traumatic Shoulder Instability: A Systematic Review and Quantitative Synthesis of the Literature. Arthroscopy : the journal of arthroscopic & related surgery : official publication of the Arthroscopy Association of North America and the International Arthroscopy Association 2016;32:944–952. 10.1016/j.arthro.2015.10.020

Marchi J, Blana D, Chadwick EK. Glenohumeral stability during a hand-positioning task in previously injured shoulders. Medical & biological engineering & computing 2014;52:251–256. 10.1007/s11517-013-1087-9

McMahon PJ, Jobe FW, Pink MM, Brault JR, Perry J. Comparative electromyographic analysis of shoulder muscles during planar motions: anterior glenohumeral instability versus normal. J Shoulder Elbow Surg 1996;5:118–123. 10.1016/s1058-2746(96)80006-1

Moroder P, Danzinger V, Maziak N, Plachel F, Pauly S, Scheibel M et al. Characteristics of functional shoulder instability. Journal of Shoulder and Elbow Surgery 2020;29:68–78. https://doi.org/10.1016/j.jse.2019.05.025

Morris AD, Kemp GJ, Frostick SP. Shoulder electromyography in multidirectional instability. Journal of shoulder and elbow surgery 2004;13:24–29.

Ogston JB, Ludewig PM. Differences in 3-Dimensional Shoulder Kinematics between Persons with Multidirectional Instability and Asymptomatic Controls. The American Journal of Sports Medicine 2007;35:1361–1370. 10.1177/0363546507300820

Pataky TC, Robinson MA, Vanrenterghem J. Region-of-interest analyses of one-dimensional biomechanical trajectories: bridging 0D and 1D theory, augmenting statistical power. PeerJ 2016;4:e2652.

Pavic R, Margetic P, Bensic M, Brnadic RL. Diagnostic value of US, MR and MR arthrography in shoulder instability. Injury 2013;44 Suppl 3:S26–32. 10.1016/s0020-1383(13)70194-3

Philp F, Faux-Nightingale A, Woolley S, de Quincey E, Pandyan A. Implications for the design of a Diagnostic Decision Support System (DDSS) to reduce time and cost to diagnosis in paediatric shoulder instability. BMC Medical Informatics and Decision Making 2021;21:78. 10.1186/s12911-021-01446-5

Philp F, Faux-Nightingale A, Woolley S, de Quincey E, Pandyan A. Evaluating the clinical decision making of physiotherapists in the assessment and management of paediatric shoulder instability. Physiotherapy 2022;115:46–57. https://doi.org/10.1016/j.physio.2021.12.330

Philp F, Freeman R, Stewart C. An international survey mapping practice and barriers for upper-limb assessments in movement analysis. Gait & Posture 2022;96:93–101. https://doi.org/10.1016/j.gaitpost.2022.05.018

Richardson RT, Rapp EA, Quinton RG, Nicholson KF, Knarr BA, Russo SA et al. Errors Associated With Utilizing Prescribed Scapular Kinematics to Estimate Unconstrained, Natural Upper Extremity Motion in Musculoskeletal Modeling. Journal of applied biomechanics 2017;33:469–473. 10.1123/jab.2016-0346

Sahrom S, Wilkie JC, Nosaka K, Blazevich AJ. Comparison of methods of derivation of the yank-time signal from the vertical ground reaction force–time signal for identification of movement-related events. Journal of Biomechanics 2021;115:110048. https://doi.org/10.1016/j.jbiomech.2020.110048

Sciascia A, Kuschinsky N, Nitz AJ, Mair SD, Uhl TL. Electromyographical comparison of four common shoulder exercises in unstable and stable shoulders. Rehabil Res Pract 2012;2012:783824. 10.1155/2012/783824

Seth A, Hicks JL, Uchida TK, Habib A, Dembia CL, Dunne JJ et al. OpenSim: Simulating musculoskeletal dynamics and neuromuscular control to study human and animal movement. PLoS computational biology 2018;14:e1006223.

Seth A, Hicks JL, Uchida TK, Habib A, Dembia CL, Dunne JJ et al. OpenSim: Simulating musculoskeletal dynamics and neuromuscular control to study human and animal movement. PLoS computational biology 2018;14:e1006223. 10.1371/journal.pcbi.1006223

Shumway-Cook A, Woollacott MH. Motor control: translating research into clinical practice: Lippincott Williams & Wilkins; 2007. (ISBN No. 0781766915)

Smith SH, Coppack RJ, van den Bogert AJ, Bennett AN, Bull AM. Review of musculoskeletal modelling in a clinical setting: Current use in rehabilitation design, surgical decision making and healthcare interventions. Clinical Biomechanics 2021;83:105292.

Spanhove V, Calders P, Berckmans K, Palmans T, Malfait F, Cools A et al. Electromyographic Muscle Activity and Three-Dimensional Scapular Kinematics in Patients With Multidirectional Shoulder Instability: A Study in the Hypermobile Type of the Ehlers-Danlos Syndrome and the Hypermobility Spectrum Disorders. Arthritis Care & Research 2022;74:833–840.

Spanhove V, Van Daele M, Van den Abeele A, Rombaut L, Castelein B, Calders P, et al. Muscle activity and scapular kinematics in individuals with multidirectional shoulder instability: A systematic review. Ann Phys Rehabil Med 2021;64:101457. 10.1016/j.rehab.2020.10.008

Stucki G. International Classification of Functioning, Disability, and Health (ICF): a promising framework and classification for rehabilitation medicine. American journal of physical medicine & rehabilitation 2005;84:733–740.

Winter DA. Biomechanics and motor control of human movement: John Wiley & Sons; 2009. (ISBN No. 0470398183)

Wu G, van der Helm FC, Veeger HE, Makhsous M, Van Roy P, Anglin C et al. ISB recommendation on definitions of joint coordinate systems of various joints for the reporting of human joint motion--Part II: shoulder, elbow, wrist and hand. J Biomech 2005;38:981–992. 10.1016/j.jbiomech.2004.05.042

Wu W, Lee PVS, Bryant AL, Galea M, Ackland DC. Subject-specific musculoskeletal modeling in the evaluation of shoulder muscle and joint function. J Biomech 2016;49:3626–3634. 10.1016/j.jbiomech.2016.09.025

