## Appendix 1 for "Young people with all forms of shoulder instability demonstrate differences in their movement and muscle activity patterns when compared to age- and sex-matched controls"

Appendix 1.Scaling marker pairs and bony landmarks used for virtual marker identification

**Scaling marker pairs used for scaling.**

| **Bone** | **X axis** | **Y axis** | **Z axis** |
| --- | --- | --- | --- |
| Thorax | IJ-C7 | PX-IJ | AC-IJ |
| Clavicle | - | - | SC-AC |
| Scapula | AC-AA | TS-AI | TS-AA |
| Humerus | ME-LE | AC-centelbow | ME-LE |
| Ulna | ME-centelbow | ME-US | ME-centelbow |
| Radius | LE-centelbow | LE-RS | LE-centelbow |

**Bony landmarks used for virtual marker identification**

- C7 spinous process (C7)
- T8 spinous process (T8)
- Insicura Jungularis (IJ)
- Processus Xiphoideus (PX)
- Articulation Sternoclavicularis (SC)
- Articulation Acromioclavicularis (AC)
- Processus Coracoideus (PC)
- Trigonum Scapulae (TS)
- Angulus Inferior (AI)
- Angulus Acromialis (AA)
- Lateral Epicondyle (LE)
- Medial Epicondyle (ME)
- Radial Styloid (RS)
- Ulnar Styloid (US)
- Styloid process of 3rd Metacarpal (MC3)
- Distal heads of the 2nd, 3rd and 5th metacarpophalangeal joints (MCP2, MCP3 and MCP5)
